# Cognitive decline heralds onset of symptomatic inherited prion disease

**DOI:** 10.1101/2020.09.14.20194217

**Authors:** Joseph Mole, Simon Mead, Peter Rudge, Akin Nihat, Mok Tzehow, John Collinge, Diana Caine

## Abstract

The clinical effectiveness of any disease-modifying treatment for prion disease, as for other neurodegenerative disorders, will depend on early treatment before damage to neural tissue is irrevocable. Thus, there is a need to identify markers which predict disease onset in healthy at-risk individuals. Whilst imaging and neurophysiological biomarkers have shown limited use in this regard, we recently reported progressive neurophysiological changes in healthy people with the inherited prion disease mutation P102L (Rudge *et al*, Brain 2019). We have also previously demonstrated a signature pattern of fronto-parietal dysfunction in mild prion disease (Caine *et al*., 2015; 2018). Here we address whether these cognitive features anticipate the onset of symptoms in a unique sample of patients with inherited prion disease. In the *cross-sectional analysis*, we analysed the performance of patients at three time points in the course of disease onset: prior to symptoms (*n* = 27), onset of subjective symptoms without positive clinical findings (*n* = 8) and symptomatic with positive clinical findings (*n* = 24). In the *longitudinal analysis*, we analysed data from twenty four patients who were presymptomatic at the time of recruitment and were followed up over a period of up to seventeen years, of whom sixteen remained healthy and eight converted to become symptomatic. In the *cross-sectional analysis*, the key finding was that, relative to a group of 25 healthy non-gene carrier controls, patients with subjective symptoms but without positive clinical findings were impaired on a smaller but very similar set of tests (Trail Making Test part A, Stroop Test, Performance IQ, gesture repetition, figure recall) to those previously found to be impaired in mild prion disease (Caine *et al*., 2015; 2018). In the *longitudinal analysis*, Trail Making Test parts A and B, Stroop test and Performance IQ scores significantly discriminated between patients who remained presymptomatic and those who converted, even before the converters reached criteria for formal diagnosis. Notably, performance on the Stroop test significantly discriminated between presymptomatic patients and converters before the onset of clinical symptoms (AUC = .83 (95% CI, 0.62-1.00), *p* =.009). Thus, we report here, for the first time, neuropsychological abnormalities in healthy patients prior to either symptom onset or clinical diagnosis of IPD. This constitutes an important component of an evolving profile of clinical and biomarker abnormalities in this crucial group for preventive medicine.

## Introduction

Pharmacological treatments for neurodegenerative disease currently focus on maintaining function, deferring cognitive or functional decline, and ameliorating symptoms such as agitation or delusions, rather than on altering the underlying pathology (Cummings and Fox, 2017). In contrast, potential disease-modifying therapy seeks to halt the underlying disease process. The clinical effectiveness of any disease-modifying treatment for prion disease, as for other neurodegenerative disorders, will to a significant extent depend on the capacity to administer treatment as early as possible, before there has been irrevocable damage to neural tissue, thereby limiting neurological injury to the greatest possible extent. That being the case, there is a pressing need to identify neuropsychological changes or biomarkers which predict future clinical diagnosis in healthy individuals at-risk of neurodegenerative diseases.

Here, we consider the human prion diseases, rare and transmissible neurodegenerative conditions which have sporadic, inherited and acquired forms and are all associated with multimeric assemblies of misfolded cellular prion protein in central nervous system and other tissues. The most common form, sporadic Creutzfeldt-Jakob disease, cannot be diagnosed prior to symptom onset, however healthy individuals at-risk of inherited prion disease (IPD) can be identified because of a diagnosis in the family and predictive genetic testing; those at-risk of acquired prion diseases can be identified based on their exposures. Ages of clinical onsets in IPD range from adolescence to the extremes of old age (Minikel *et al*., 2019) making it difficult to design preventive clinical trials. To date, imaging and neurophysiological biomarkers have shown limited use in prediction of clinical onset of IPD (Franko *et al*., 2016; Zanusso *et al*., 2016) but we recently reported progressive neurophysiological changes in healthy people with the IPD mutation P102L (Rudge *et al*., 2019).

Notwithstanding both the variety of causes of human prion diseases and their phenotypic heterogeneity, we have previously demonstrated in a large mixed cohort that early disease is characterised by a signature pattern of cognitive deficits (Caine *et al*., 2015; 2018). These comprise prominent executive impairment, parietal dysfunction, dynamic aphasia, and reduced motor speed. We argued further that inasmuch as the cognitive deficits arise in the context of a movement disorder (most often ataxia and/or myoclonus), prion disease should be considered in the constellation of movement disorders with associated dementia syndromes including corticobasal degeneration, progressive supranuclear palsy, Lewy body disease and Huntington’s disease (HD).

If the pattern of cognitive deficits is useful for diagnosis in the early stages of disease, might they indeed herald the onset of symptoms in patients with IPD? This question has previously been addressed in the context of Huntington’s disease which, like IPD, provides the opportunity to observe gene-positive individuals prior to and during conversion to disease onset. In a study of 136 asymptomatic patients at risk of HD, to whom an extensive battery of cognitive tests was administered, Snowden *et al*. (2002) found only a mildly reduced performance on undemanding psychomotor tasks in participants with the HD mutation compared to those without. They argued that this represented subtle cognitive decline in pre-clinical subjects, relative to the very marked cognitive decline that is a feature of early disease, suggesting more precipitous decline in cognitive ability around clinical onset. Brandt *et al*. (2008) found reduced performance only on the Wisconsin Card Sorting Test amongst ‘converters’ who were closer to disease onset. On the other hand, Martinez-Horta *et al*. (2016) reported that neuropsychiatric symptoms are highly prevalent even in the pre-manifest stage of HD, with increasing prevalence of irritability, apathy and executive dysfunction closer to onset. This question has never previously been addressed in the context of prion disease.

In a study comprising near comprehensive nationwide recruitment of patients with all types of prion disease over more than ten years, we have had the opportunity to evaluate the cognitive profiles of a large cohort of patients with, and at risk for, IPD (Thompson *et al*., 2013). This has included patients identified as gene-positive at recruitment, some of whom have, over the course of the study, converted to become affected by the disease. Thus we have been uniquely placed to investigate and identify the first markers of disease onset including, importantly, the earliest changes in cognitive status. In addressing the question of whether cognitive change might herald disease onset in these patients we have adopted two strategies. The first is a cross-sectional comparison of patients at three different moments in the course of disease onset: prior to symptoms occurring; when subjective symptoms arise but without positive clinical findings; and symptomatic with positive clinical findings resulting in firm diagnosis. The second is a longitudinal study of patients who were presymptomatic at the time of recruitment but who converted to become symptomatic and were later diagnosed with the disease during the course of the study.

## Materials and methods

### Participants

Patients were recruited through the NHS National Prion Clinic (NPC) at the National Hospital for Neurology & Neurosurgery, UCLH NHS Foundation Trust, London, U.K. Longitudinal study of patients included repeated annual neurological investigations and neuropsychological assessments until they showed signs of becoming symptomatic, when monitoring became more frequent, 6 monthly or more often as required. 804 patients with suspected or confirmed prion disease were recruited to the National Prion Cohort Monitoring Study between 2008 and 2019^1^. Many of these were already symptomatic at the time of recruitment, indeed most were too severely impaired to undergo detailed cognitive assessment.

Clinical phenotype of IPD is highly variable with many cases, typified by the E200K mutation, showing a rapidly progressive dementia with ataxia and myoclonus indistinguishable from sporadic Creutzfeldt-Jakob disease. In other forms however, typified by the P102L mutation and the Gerstmann-Straussler-Scheinker clinical syndrome, disease onset is usually insidious with gradually progressive motor, neuropsychological, or neuropsychiatric deficits which can also be obscured by co-existing depressive illness or other fluctuating symptoms which are common in at-risk groups (Thompson *et al*., 2013). As a result, disease onset can itself be considered to occur over a period of time during which different stages can be identified. The earliest is that of the *first symptom* experienced by the patient that subsequently progresses to form part of the syndrome of IPD. Such symptoms gradually become more *significant* causing clear objective disability. Usually, between these two stages, a *clinical diagnosis* of IPD will be made, based on the confirmation of subjective symptoms with positive test results or examination findings. In this study the *symptom onset* was defined retrospectively by the clinical team as the date of onset of symptoms that subsequently progressed into the IPD clinical syndrome. *Clinical diagnosis* was defined as the date at which the clinical team were confident enough, based on the totality of the clinical assessment and investigations, to feed-back to the patient that IPD was the cause of symptoms. The development of *significant symptoms* was defined as the date at which an abnormality in activities of daily living was first documented, aided by use of the Medical Research Council Prion Disease Rating Scale (MRC Scale), a measure of overall disease severity comprising neurological, cognitive, and functional components (Thompson *et al*., 2013). Patients were eligible for the *cross-sectional analysis* if they had IPD and had completed a neuropsychological assessment and the MRC scale. Patients were assigned to three groups: *presymptomatic* (all had MRC Scale score = 20/20, *n* = 27), *symptom onset* with MRC Scale score = 20 (*n* = 8), and *significant symptoms* with MRC scale score < 20 (*n* = 24). Some of the patients in the latter two groups were reported in our original study of cognition in prion disease (Caine *et al*., 2015). Patients’ scores were compared with those of a matched control group of 25 healthy non gene-carriers, recruited from amongst the patients’ families, to control for possible confounding factors such as education and IQ, who also underwent repeated assessment.

For the *longitudinal analysis*, the patients of particular interest were those with a known prion mutation who were presymptomatic at recruitment but who converted to be diagnosed with the disease during the course of the study (‘converters’). Patients were included in this analysis if they met this criterion and had undergone at least two neuropsychological assessments *prior to becoming symptomatic and prior to clinical diagnosis*. These time points of *onset of first clinical symptoms* and formal *clinical diagnosis* being rendered to the patient were chosen as the salient moments of onset for this component of the study.

There were eight converters (genetic group: P102L, *n* = 5; 5-octapeptide repeat insertion (OPRI), *n* = 1; 6-0PRI, *n* = 1; D178N, *n* = 1). Their clinical symptoms were heterogeneous: for some patients their first symptoms were cognitive, whereas for others they were physical (see Supplementary Table 1). For this analysis, the control group comprised participants who had tested positive for one or another prion protein gene mutation, but who remained presymptomatic (*n* = 16) and had undergone at least two neuropsychological assessments. At the time of the analysis, this group had remained presymptomatic for an average of 2.88 *(SD* = 1.96) years after their last assessment.

Ethics approval for the study was granted by the Eastern Multicentre Research Ethics Committee. The research was done in accordance with the Declaration of Helsinki and informed consent was obtained from patients.

### Investigations

In addition to systematic neurological examination, all participants underwent neuropsychological investigation comprising a comprehensive battery of standardized tests selected originally to characterise the cognitive deficits in prion disease (Cordery *et al*., 2005; Caine *et al*., 2015). It included the following: premorbid optimal level of function (National Adult Reading Test (Nelson, 1982)) current intellectual functioning (WAIS-III (Wechsler, 1997)); visual perception and visuospatial function (Visual Object and Space Perception Battery, Object Decision, Cube Analysis (Warrington and James, 1991)); (The Adult Memory and Information Processing Battery, Figure copy (Coughlan and Hollows, 1986)); spelling (Graded Difficulty Spelling Test (Baxter and Warrington, 1994)); calculation (Graded Difficulty Arithmetic Test (Jackson and Warrington, 1986)); limb praxis [meaningless gesture repetition (Goldenberg, 1996)]; visual and verbal recognition memory (Recognition Memory Test (Warrington, 1984)); language, including category (‘Animal’) fluency (Spreen and Strauss, 1998); object naming (Graded Naming Test (McKenna and Warrington, 1980)); synonym matching (Warrington *et al*., 1998); and sentence comprehension (Test for Reception of Grammar (Bishop, 1989)); executive function (Stroop Test (Trenerry *et al*., 1989); verbal fluency (Spreen and Strauss, 1998); Trail Making Test part B (Halstead and Reitan, 1985)) and information processing speed (Trail Making Test part A (Halstead and Reitan, 1985). Levels of anxiety and depression were measured using the Hospital Anxiety and Depression Scale, an objective measure of mood disturbance (Zigmond and Snaith, 1983). The tests selected for this study were all those on which there was a comprehensive longitudinal set of data for all or most of the participants. For participants included in the longitudinal analysis, on average, the converter group completed 6.50 (SD = 1.51) assessments and the presymptomatic gene-positive control group completed 3.91 *(SD* = 2.77) assessments.

### Statistical analyses

Statistical analyses were carried out using IBM SPSS Statistics 26. Skewness and kurtosis was assessed, by inspecting boxplots, and homogeneity of variances was assessed, using Levene’s test. All variables were continuous, with the exception of gender, which was a dichotomous variable. If data were not normally distributed, and this could not be corrected by transformation, non-parametric statistics were used (Mann–Whitney U-tests).

In the *cross-sectional analysis*, we first assessed whether the three patient groups *(presymptomatic* MRC = 20; *symptom onset* MRC = 20; and *significant symptoms* MRC < 20) and healthy controls differed in terms of demographic variables (age, number of years of education and premorbid intelligence). Next, the three patient groups’ scores on neuropsychological tests and the Hospital Anxiety and Depression Scale at the time of first assessment were compared with those of the healthy control group. Comparisons were made using Independent t tests or Mann–Whitney U tests. The Bonferroni correction was applied to all analyses and only results that remained significant after adjusting for multiple comparisons (i.e. *p* < .017) are reported.

Before undertaking the *longitudinal analyses* we sought to determine whether patients and presymptomatic gene-positive controls differed in terms of demographic variables (age, number of years of education and premorbid intelligence), performance on neuropsychological tests and mood at the time of their first assessment. Comparisons were made using Independent t tests or Mann–Whitney U tests. Gender differences were investigated using Chi-square goodness-of-fit tests.

The *longitudinal analyses* comprised three components. The first analysis sought to determine whether the converters declined relative to presymptomatic gene-positive controls on neuropsychological tests and mood prior to: 1) confirmation of symptom onset, and 2) confirmation of disease onset, as defined above. Analysis of variance (ANOVA) was conducted, with subject group as the between-subjects factor (symptomatic patients vs. presymptomatic gene-positive controls) and time of assessment as the within-subjects factor. In the first analysis, the within-subjects factors were first vs. last assessment before symptom onset and in the second analysis, they were first vs. last assessment before confirmation of disease onset. Missing data were managed conservatively: where data were missing from individual tests at the time of the first assessment, data from the next assessment on which the test was completed were inputted and treated as the initial datum for that test (9.18% of first assessment scores). Data were missing from the last assessment before symptom onset in only one instance (0.21% of last assessment scores), where tests were not administered due to time constraints. Data were missing from the last assessment before confirmation of disease onset in only two instances (0.43% of last assessment scores), where a converter had not been administered visual and verbal recognition memory tests, as they were already found to be impaired on these tests in the previous assessment. In these instances, scores from the previous assessment were inputted and treated as the final datum for that test.

The second analysis used receiver operating characteristics (ROC) to determine whether it was possible to discriminate between converters and presymptomatic gene-positive controls based on extent of decline in neuropsychological performance and mood. Two analyses were undertaken. In the first, the independent variable was the difference in raw scores between the first and last assessment before symptom onset. In the second, the independent variable was the difference in raw scores between the first and last assessment before confirmation of disease onset. Only data from tests where the first longitudinal analysis demonstrated a significant interaction between group and time of assessment were analysed.

The third analysis aimed to investigate how cognition declined over time in the converter group vs the presymptomatic gene-positive control group. As before, only data from tests where the first longitudinal analysis demonstrated a significant interaction between group and time of assessment were analysed. The independent variable for this analysis was the raw scores on each of the neuropsychological tests analysed. For these tests data were extracted from neuropsychological assessments that occurred within the following four time-points: time-point one (0-12 months before diagnosis), time-point two (13-24 months before diagnosis), time-point three (25-36 months before diagnosis) and time-point four (37-48 months before diagnosis). In the presymptomatic gene-positive control group, data were extracted for time-point four from the last assessment undertaken. For time-points two, three and four, data were extracted from assessments conducted one, two and three years prior to that. Repeated measurements of the same psychological test over time were analysed using a linear mixed model approach including allowing for individual random effects in the rate of change in the test score (Stata 15.0, StataCorp).

Again, missing data were inputted conservatively. The ‘last observation carried forward’ method was used. In two cases in the converter group and six cases in the presymptomatic gene-positive control group, patients did not have a neuropsychological assessment during one of the four time-points (e.g. 13-24 months before diagnosis). In these cases data were entered from the previous time-point (e.g. 25-36 months before diagnosis).

## Results

### Cross-Sectional analysis

#### Descriptive characteristics

Descriptive characteristics and scores on neuropsychological tests and the Hospital Anxiety and Depression Scale at the time of the first assessment are presented in Supplementary Table 2. The presymptomatic MRC = 20, symptomatic MRC = 20 or symptomatic MRC < 20 groups did not significantly differ from the healthy control groups (i.e. *p* > 0.017) in terms of sex, age, education and pre-morbid intelligence.

#### Cognitive performance

In comparison to the control group of healthy non gene-carriers, the performance of the presymptomatic MRC = 20 group was intact on all tests apart from the Graded Difficulty Arithmetic Test (t (31) = −2.570, *p* = .008; see Supplementary Table 2), likely reflecting the very high working memory demands of this oral task. In contrast, patients in the symptomatic MRC = 20 group had significantly weaker *Performance IQ* than healthy controls (t (31) = −2.487, *p* = .009) and significantly weaker scores on tests of *praxis* (meaningless gesture repetition, right hand; *U* = 39.50, *z* = −3.282, *p* = .0165), *delayed non-verbal recall memory* (AMIPB Figure, delayed recall; *U* = 40.00, *z* = −2.522, *p* = .005), *executive function* (Stroop Test: *U* = 32.00, *z* = −2.537, *p* = .005) and *information processing speed* (Trail Making Test, part A: *U* = 38.50, *z* = −2.588, *p* = .004). Patients in the symptomatic MRC <20 group had significantly weaker performance than healthy controls on *all tests*, with the exception of recognition memory and synonym matching (all *p* <.017).

#### Mood

There were no significant differences in levels of anxiety and depression between the healthy controls and any of the patient groups included in the cross-sectional analysis.

### Longitudinal analysis

Descriptive characteristics and baseline neuropsychological tests

Descriptive characteristics and scores on neuropsychological tests and the Hospital Anxiety and Depression Scale at the time of the first assessment are presented in Supplementary Table 3. Converter and presymptomatic gene-positive control groups were matched (i.e. *p* > 0.05) for sex, age, education and pre-morbid intelligence. Moreover, there was no significant difference between converters (Mea*n* = 85.75, *SD* = 44.49, range = 34 - 188) and presymptomatic gene-positive controls (Mea*n* = 82.25, *SD* = 55.91, range = 12 - 196) in terms of the number of months between their first and last assessment, *t* (22) = .971, *p* = .879.

> *(i) Comparison between converters and presymptomatic gene-positive controls on cognitive tasks and mood*

The first analysis sought to determine whether the converters declined relative to presymptomatic gene-positive controls on neuropsychological tests prior to: 1) symptom onset, and 2) confirmation of disease onset. In the first of these analyses, the converters showed a significantly greater decline in Stroop test performance between the first and last assessment before symptom onset, relative to that of the presymptomatic gene-positive controls, as shown by a significant interaction between group and time of assessment (*F* (1, 22) = 5.71, *p* = .026). No significant interactions between group and time of assessment were found for performance on any other neuropsychological tests or for levels of anxiety and depression. In the second of these analyses, the converters’ Performance IQ but not Verbal IQ declined between the first and last assessment before confirmation of disease onset, relative to presymptomatic gene-positive controls, as shown by a significant interaction between group and time of assessment (*F* (1, 22) = 13.95, *p* = .001). On tests of executive function and information processing speed, the converters also showed a decline in performance between the first and last assessment before confirmation of disease onset, relative to presymptomatic gene-positive controls, as shown by a significant interaction between group and time of assessment as follows: Stroop Test (*F* (1, 22) = 28.15*,p* < .001); Trail Making Test, parts A and B (*F* (1, 22) = 10.18, *p* = .004; *F* (1, 22) = 9.63, *p* = .005, respectively). The only exception was a non-significant trend towards an interaction between group and time of assessment for phonemic fluency performance (*F* (1, 22) = 3.868, *p* = .062). Notably, while there was no significant difference between the groups at the time of the first assessment, the converters’ phonemic fluency performance was significantly poorer than that of presymptomatic gene-positive controls at the last assessment before diagnosis, *t* (22) = 1.948, *p* = .032. Mixed ANOVAs revealed no significant interaction between group (converters vs. presymptomatic gene-positive controls) and time of assessment (first vs. last assessment before time of confirmation of disease onset) in terms of performance on neuropsychological tests in the domains of perception, limb praxis, spelling, calculation, visuo-construction, verbal and non-verbal recognition memory, or language.

The converters showed an increase in Hospital Anxiety and Depression Scale anxiety scores between the first and last assessment before confirmation of disease onset, relative to presymptomatic gene-positive controls, as shown by a significant interaction between group and time of assessment (*F* (1, 14) = 22.53, *p* = .047). Qualitatively, between the first and last assessment, average anxiety scores increased from the normal range to the ‘doubtful case’ range (Zigmond and Snaith, 1983). There was no significant interaction between group and time of assessment for Hospital Anxiety and Depression Scale depression scores.

> *(ii) ROC analysis of converters vs presymptomatic gene-positive controls*

A ROC curve analysis was undertaken to determine whether it was possible to discriminate between converters and presymptomatic gene-positive controls based on extent of decline in neuropsychological performance and mood. This was undertaken for the measures on which the previous analysis showed a significant interaction. In the first analysis, decline in Stroop test scores between the first assessment and last assessment before symptom onset significantly discriminated between converters and presymptomatic gene-positive patients (AUC) = .83 (95% CI, 0.62-1.00), *p* =.009. No other independent variables significantly discriminated between the groups at this stage. In the second analysis of decline in performance between the first assessment and last assessment before confirmation of disease onset, decline in Performance IQ, Stroop test, and Trail Making Test, parts A and B scores significantly discriminated between converters and presymptomatic gene-positive patients (see Figure 1). Discriminability was excellent for the Stroop test (area under the curve (AUC) = .95 (95% CI, 0.86-1.00), *p* <.001) and good for Performance IQ (AUC = .87 (95% CI, 0.71-1.00), *p* =.004) and the Trail Making Test, parts A and B (AUC = .81 (95% CI, 0.60-1.00), *p* =. 016, AUC = .84 (95% CI, .63-100), *p* =.008, respectively). Change in Hospital Anxiety and Depression Scale anxiety scores did not significantly discriminate between the groups. Because of this, levels of anxiety and depression were not investigated in the following analysis.

**Figure 1.**
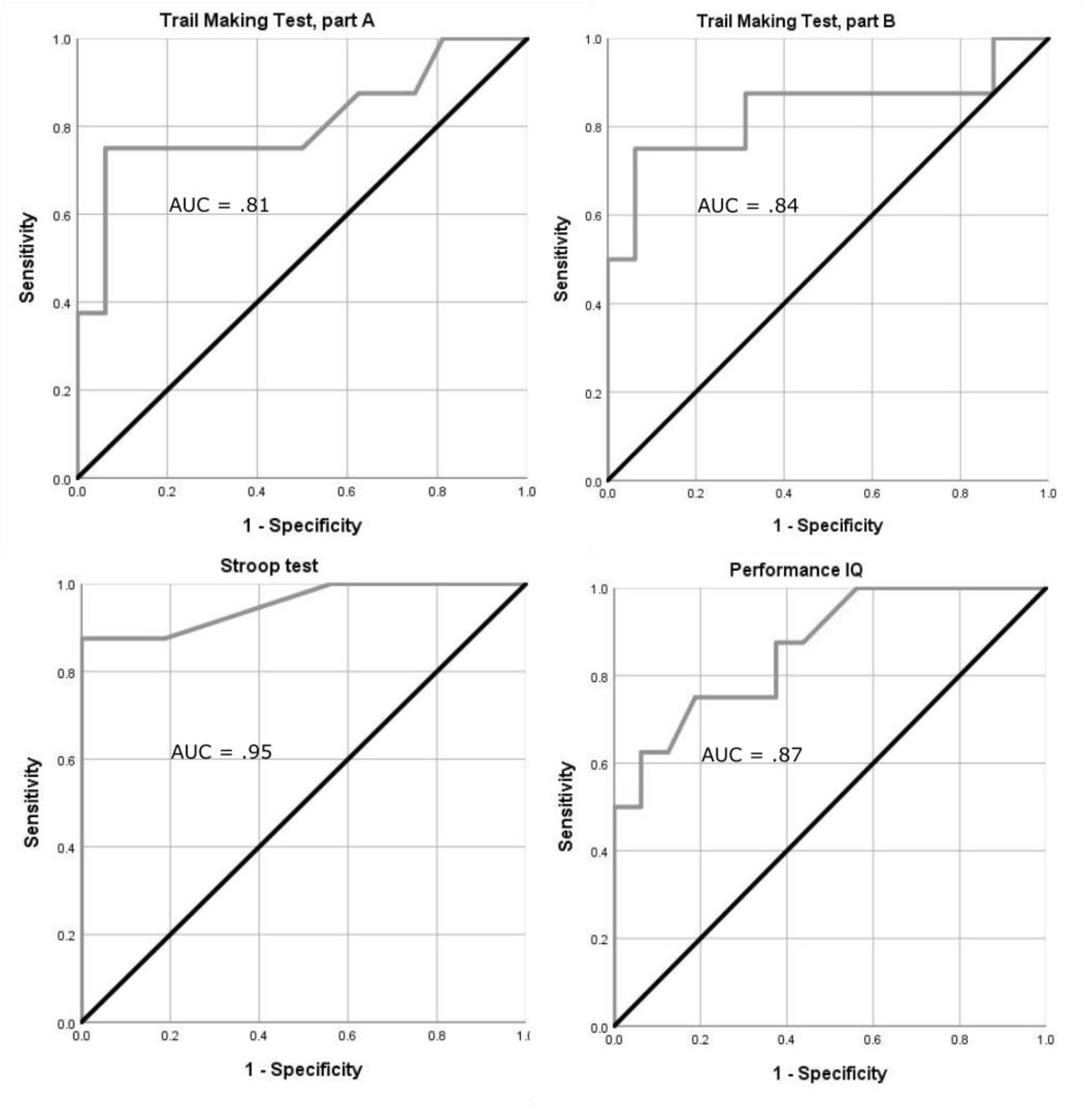
ROC curve analysis of converters vs. asymptomatic gene-positive controls.

> *(iii) Cognitive decline over time*

In this analysis we sought to define a rate of change in test score in converters and presymptomatic gene-positive controls, over a four year time period prior to diagnosis, using a linear mixed model approach (see methods). The TMTA showed a change of +3.8s per year in converters compared with presymptomatic gene-positive controls (coefficient 3.8s (95% CI 1.3 to 6.2s), P=0.003), the TMTB a change of +22.5s per year (coefficient 22.5 (95% CI 12.2 to 32.9), P<0.001), the Stroop test a change in score of −10.8 per year (coefficient −10.8 (95% CI −14.9 to −6.8), P<0.001), Performance IQ a change in score of −4.3 per year (coefficient −4.3 (95% CI −7.9 to −0.77), P=0.017 (see Figures 2-5).

**Figure 2:**
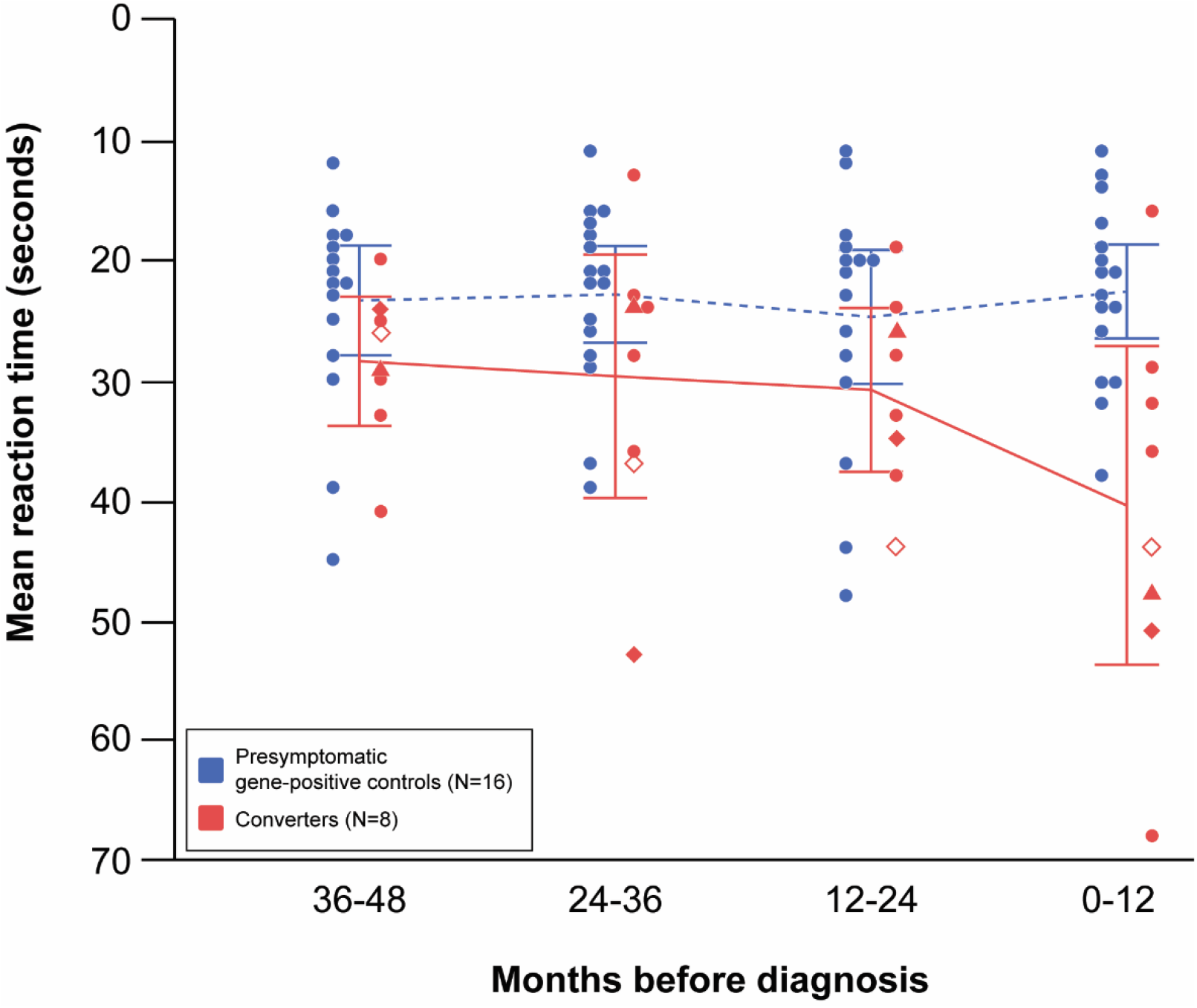
Trail Making Test, Part A.

**Figure 3:**
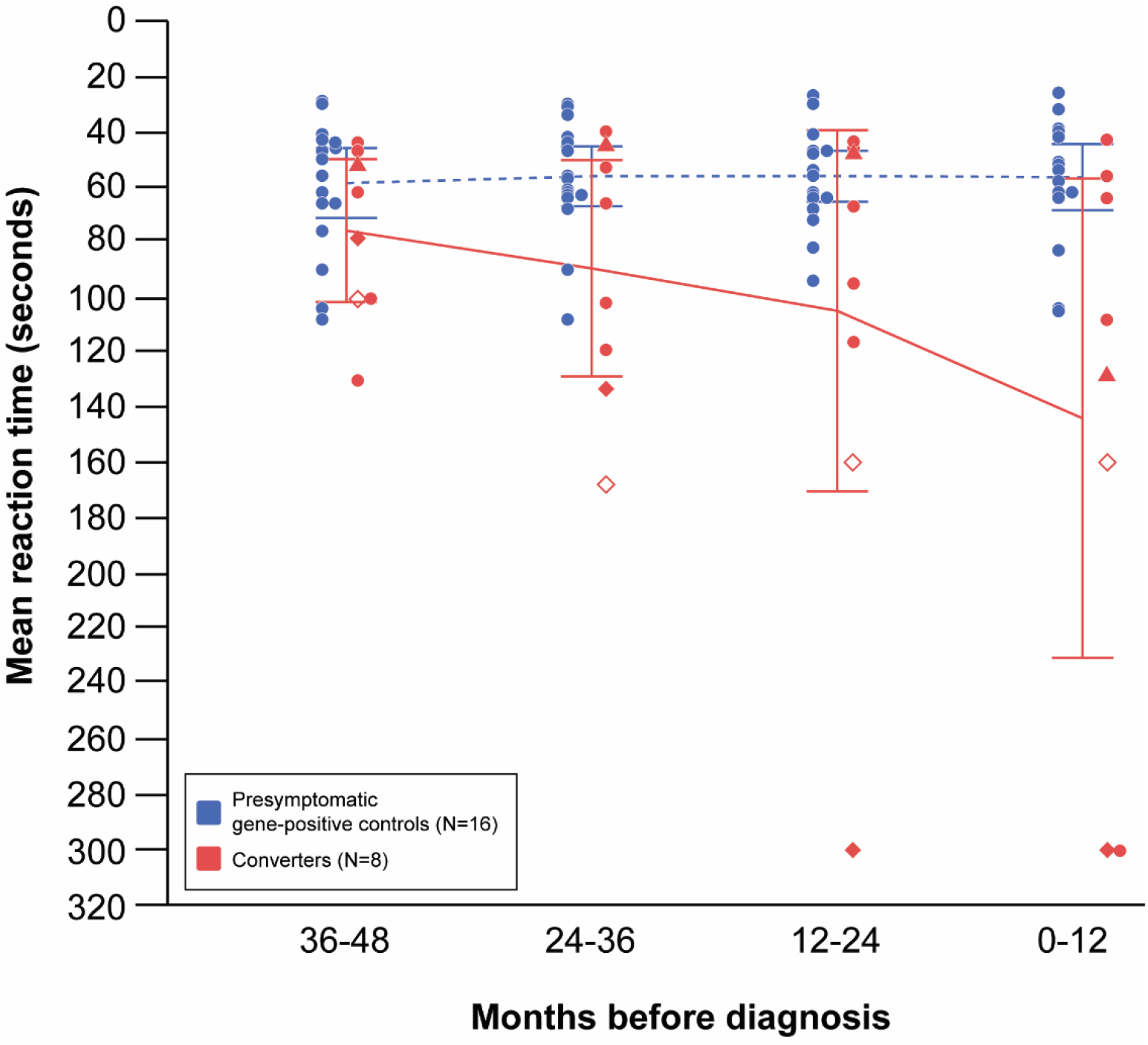
Trail Making Test, Part B.

**Figure 4:**
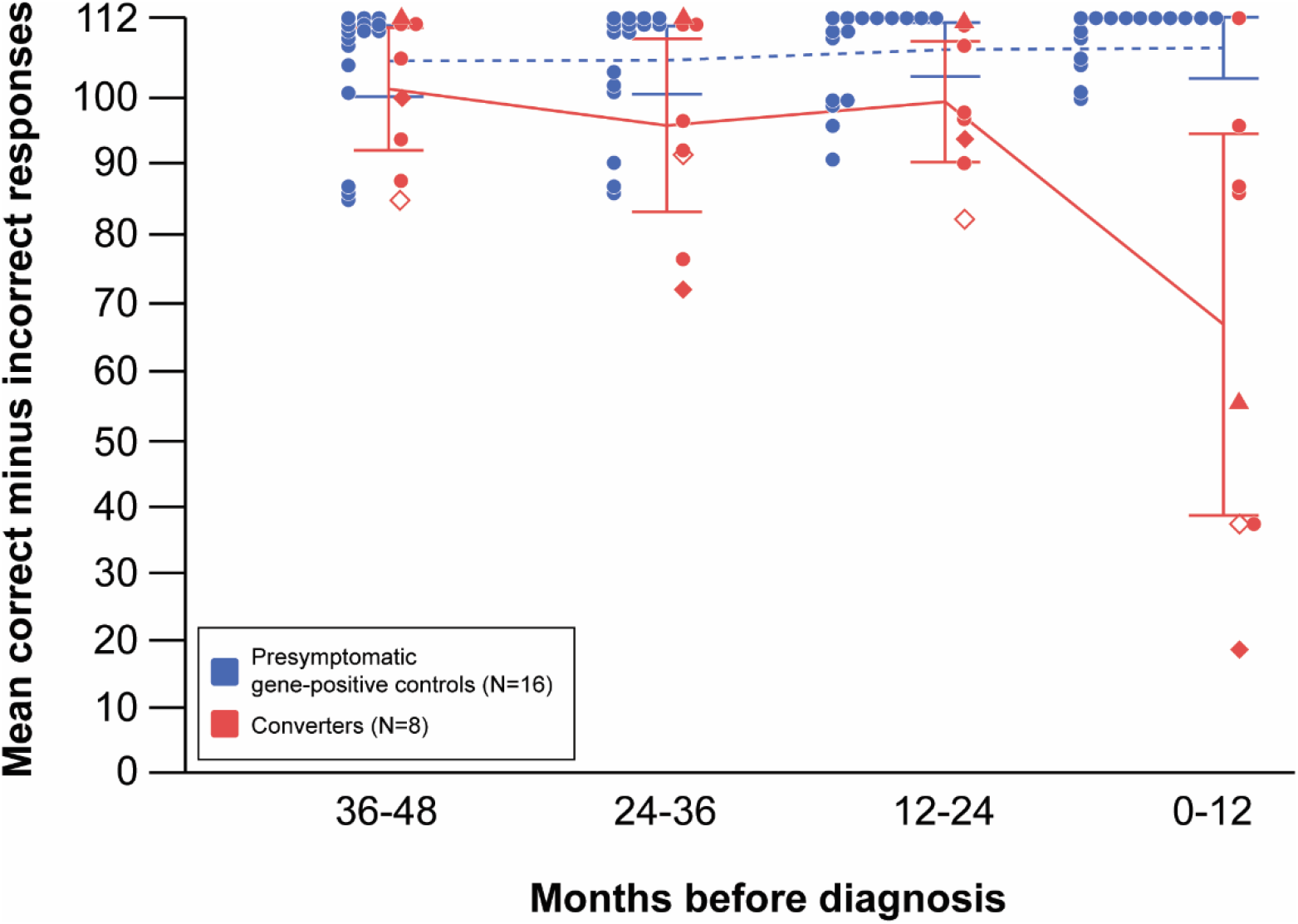
Stroop test.

**Figure 5:**
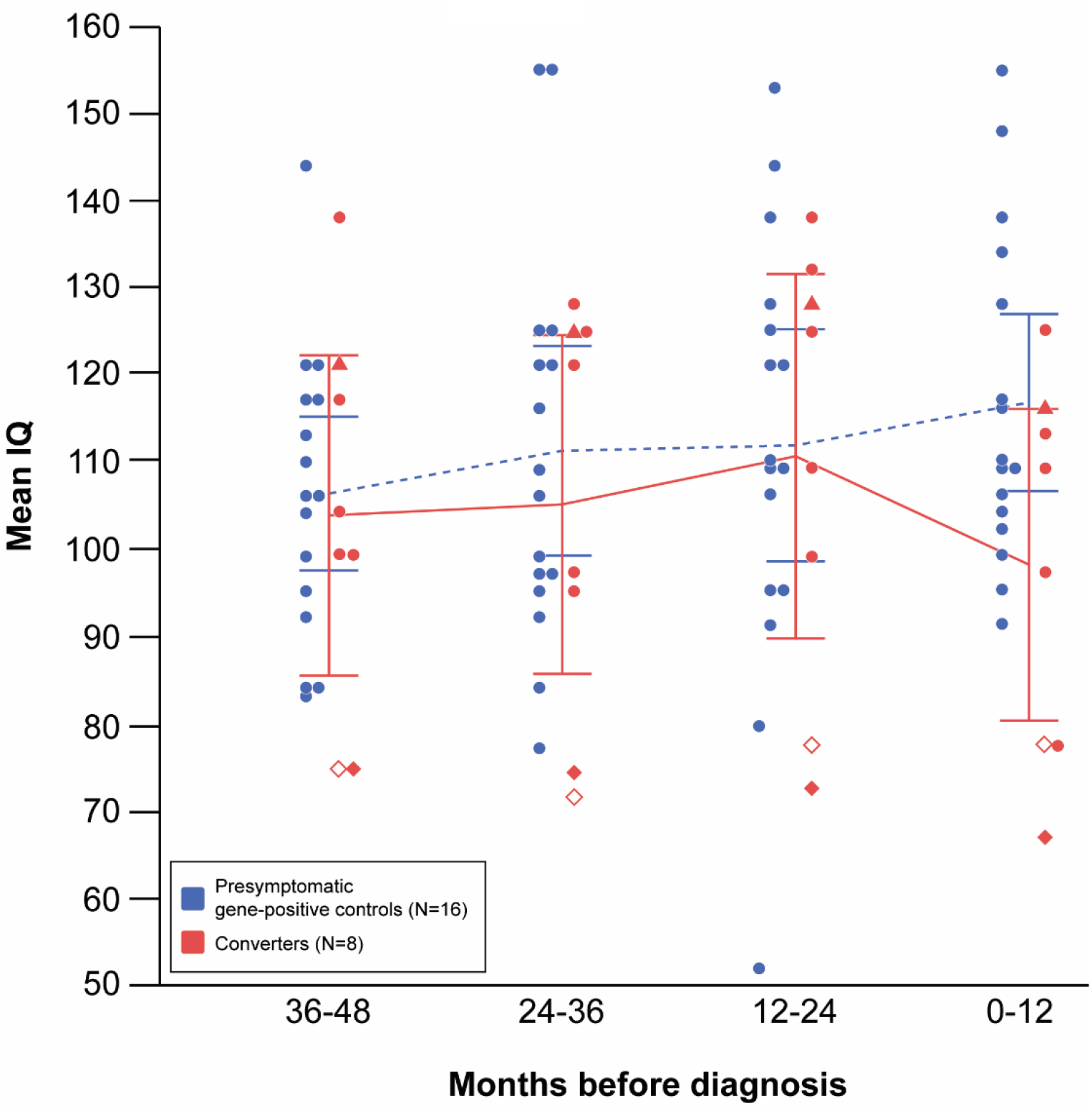
Performance IQ.

## Discussion

Prion disease is characterised by an apparently heterogeneous complex of neurological and neuropsychological signs and symptoms. If potential future disease modifying treatments are to be effective they will need to be instituted at the earliest possible opportunity in order to prevent any or further brain damage. We do now know, for instance, that neurophysiological changes can be detected in healthy carriers of the prion mutation P102L (Rudge *et al*, Brain 2019). The timing of onset of cognitive change relative to disease onset has never previously been investigated in this context. Overall the findings of this study suggest that subtle but definitive cognitive change can also be demonstrated in gene carriers prior to onset of the first clinical symptoms of the disease and confirmation of disease onset.

As we have shown previously, and has been demonstrated elsewhere (see, for example, Cordery *et al*., 2005), symptomatic prion disease patients experience pervasive cognitive decline as seen in our cross-sectional study. Previously we demonstrated that a coherent constellation of cognitive variables associated with fronto-parietal function are the leading cognitive features in mild prion disease. In this study, in which we have been able to examine evolving cognitive profiles over time, we have repeated and refined that finding. The key factor in the present work is that scores on a smaller set of tests are evidently impaired prior to the patient reaching criteria for formal diagnosis of the disease, whether comparison is made cross-sectionally or longitudinally.

*Cross-sectional analysis* revealed that patients experiencing symptoms but still functionally independent for activities of daily living (the symptomatic MRC = 20 group) performed less well than healthy non-gene carrier controls on a similar group of tests, namely *praxis, delayed non-verbal recall memory, Performance IQ, executive function* and *information processing speed*. This clearly demonstrates that specific cognitive changes are present at the earliest stages of disease onset, prior to significant symptoms.

The *longitudinal study* of gene carriers similarly demonstrated that, prior to confirmation of disease onset, converters - patients who were presymptomatic at recruitment but who developed the disease while under investigation-significantly declined relative to presymptomatic gene-positive controls in their Performance IQ and on tests of executive function and information processing speed. Strikingly, converters performed significantly less well on one test of executive function, the Stroop test, prior to onset of the first clinical symptoms of the disease. The second phase of the analysis also showed that converters could be discriminated from presymptomatic gene carriers on the same set of tests, prior to the onset of the first clinical symptoms (Stroop) and confirmation of disease onset (Performance IQ, Stroop, and TMT parts A & B). Finally, by analysing time series data up to four years prior to clinical diagnosis we show evidence of a linear deterioration in four neuropsychological tests. In contrast with similar studies of patients with HD, another genetically determined neurodegenerative disease that compromises both movement and cognition (Snowden et al., 2002; Brandt et al., 2008), cognitive change was found here clearly to herald onset of disease.

The findings are particularly striking because the converters were not homogenous with respect to gene mutation: five of the eight converters had the P102L mutation, while the others each had a different mutation (5-OPRI, 6-OPRI, D178N: see supplementary Table 1). P102L is a genetic form in which considerable heterogeneity has been observed but in which onset is often characterised by a slowly progressive cerebellar ataxia with later onset cognitive impairment, sometimes the reverse (Webb et al, Brain 2008), while 6-OPRI is known to present with predominantly cognitive decline. Thus, both the heterogeneity of the group, and the fact that some of the patients had predominantly motor onset (see Supplementary Table 1), underscore the robustness of the group finding. The group of ‘converter’ patients was necessarily small. This is unsurprising in view of the rarity of the condition and the difficulties attendant on consistent long-term follow-up. Given these limitations, our findings must remain tentative. Prospective testing in an independent group of patients based on the experiences in other countries will be necessary to enable more confidence in their properties and generalisability.

The National Prion Clinic has now been recruiting patients for 12 years. At the start of the study the Trail Making Test part B and the Stroop Test were both considered to be reliable tests of executive function. Recent intensive investigation of these instruments has shown that the TMT B is in fact a robust test for detection of brain dysfunction (Chan et al., 2015) but with limited specificity for frontal lesions, while the Stroop Test has been confirmed to be consistently significantly impaired in patients with focal lesions of the prefrontal cortex (Cipolotti et al., 2016). Thus, failure on these two tasks, which we have now demonstrated in a variety of ways both in the current and previous study (Caine et al. 2015), to be present early in prion disease, accords with the facts both that the brain pathology in prion patients is not strictly speaking focal and that executive dysfunction tends to be a very early feature of the disease. Both tasks are also timed, as are the sub-tests comprising the Performance IQ scale. Abnormal performance on the TMT A amongst the converters clearly indicates reduced processing speed which may, of course, also have some impact on performance on other tasks.

It is noteworthy that for performance on the only executive test that was not significant in the longitudinal analysis (phonemic fluency) there was a non-significant trend for an interaction between group and time of assessment. Moreover, while the interaction effect did not reach significance, converters’ phonemic fluency performance was significantly poorer than that of presymptomatic gene-positive controls at the last assessment before diagnosis. We have previously found that, in a larger sample of 40 patients with prion disease, who were symptomatic but nevertheless still able to undergo detailed language testing, phonemic fluency was impaired in 70% of patients (Caine et al., 2018). Moreover, performance was strongly correlated with performance on two of the executive tests that were significant in the longitudinal analyses (Stroop test, Trail Making Test part B). Thus, while the effect may have been too subtle to confirm in the current sample, it is plausible that declining phonemic fluency comprises part of the emerging neuropsychological profile of conversion in IPD.

In addition to cognitive decline, the longitudinal analysis revealed a slight increase in levels of anxiety in converters, although the extent of this increase did not significantly discriminate this group from asymptomatic gene-positive controls. It is likely that this is because anxiety levels were relatively low, with the average scores in the converter group not exceeding the ‘doubtful case’ range. Anxiety is therefore not thought significantly to have impacted the neurosychological findings although this question clearly warrants further investigation to be confident of this interpretation.

The MRC Prion Disease Rating Scale (Thompson et al. 2013) was designed to capture change over time in prion patients whose course is sometimes, especially in the case of sporadic disease, very rapid. While its psychometric properties are robust, and while it includes items on cognitive capacity, the latter is evaluated by rater judgement on only four items (memory and orientation, speech, judgement and problem-solving tool use). It is therefore not surprising that at the point in time that was defined as being the point of disease onset in the converters, all of them produced maximum scores on the MRC scale. Conversely, at that point the converters were starting to show signs of cognitive decline on more searching tests. Once disease was formally diagnosed, the MRC scale addressing ADL’s, motor function and cognition has been shown to be able sensitively to capture change over time (Thompson et al., 2013). Prediction of future clinical diagnosis in at-risk individuals is a highly sensitive matter. A clinical tool should not be used without confidence in how well it works, or an understanding of its purpose. Some at-risk individuals would prefer that a diagnosis of IPD is considered only when it is essential for clinical care. Others however, seek as much information as possible about their condition, believing that information that helps predict the future will help them make plans for their care, financial and personal affairs whilst they have the ability to do so. Others again are motivated by the altruistic desire to improve clinical trial methods such that disease-modifying therapeutics that prevent or delay symptom onset can be tested with more robust outcomes and definitions, and developers see the potential for an effective test of their compounds. Whichever may be the case, here we describe work with a special if small group of patients who developed IPD during longitudinal neuropsychological study, which will be invaluable in the progress towards development of a neuropsychological test battery that can predict future diagnosis of IPD and can detect the first clinical symptoms of the disease.

We report for the first time neuropsychological abnormalities in healthy patients before symptom onset and clinical diagnosis of IPD, an important component of an evolving profile of clinical and biomarker abnormalities in this crucial group for preventive medicine.

## Data Availability

The data that support the findings of this study are available from the corresponding author, DC, upon reasonable request.

## Funding information

The Cohort study was funded by the Department of Health (England) and the UCLH/UCL Biomedical Research Centre with additional support from the Medical Research Council. SM and JC are National Institute of Health Research Senior Investigators. Tze How Mok is supported by the Alzheimer’s Society UK. JM is supported by the National Brain Appeal.

## Competing interests

The authors have no competing interests to declare.

## Acknowledgements

We thank patients, and other participants of the Cohort study, for their generous contribution of time and effort. We also thank past and present colleagues at the NPC. The Cohort study was funded by the Department of Health (England) and the National Institute of Health Research’s (NIHR) Biomedical Research Centre at University College London Hospitals NHS Foundation Trust. Simon Mead and John Collinge are NIHR Senior Investigators. Some funding was provided by the Medical Research Council (UK). JM is funded by the National Brain Appeal.

## Conflict of Interest

None declared.

## Declaration of authorship

J. Mole: Conceptualization, Formal analysis, Methodology, Investigation, Writing - original draft, Writing - review & editing. S. Mead: Conceptualization, Formal analysis, Funding acquisition, Methodology, Investigation, Writing - review & editing, Supervision. P. Rudge, Conceptualization, Methodology, Writing - review & editing, Supervision. A. Nihat: Methodology, Writing - review & editing. M. Tzehow: Methodology, Writing - review & editing. J. Collinge: Funding acquisition, Writing - review & editing, Supervision. D. Caine: Conceptualization, Methodology, Investigation, Writing - original draft, Writing - review & editing, Supervision.

## Footnotes

1 In some cases, patients had undergone some of their neuropsychological assessments prior to enrolment in the study, as part of their routine clinical care, but gave their consent for previously collected data to be analysed retrospectively. This allowed us to analyse changes in cognitive function over a period of up to seventeen years

## Notes

### Competing Interest Statement

The authors have declared no competing interest.

### Author Declarations

Ethics approval for the study was granted by the Eastern Multicentre Research Ethics Committee.

